# Maternal Cigarette Smoking Trajectories and Risk of Low Birthweight in the United States: A Population-Based Analysis of 2023 to 2024 Natality Data

**DOI:** 10.64898/2026.10.01.26364505

**Authors:** Chika Onwuameze, Godyson Orji, Martin Ayuk Bate, Vivian Ogechi Madu, Philip Kaya, Joy Inalegwu

## Abstract

**Background:** Maternal cigarette smoking is associated with adverse birth outcomes, but low birthweight may reflect preterm delivery, restricted fetal growth, or both. Contemporary evidence distinguishing smoking trajectories is limited.

**Objectives:** To examine associations between maternal cigarette-smoking trajectories and low birthweight among U.S.-resident singleton live births in 2023 to 2024.

**Methods:** We analysed 5,861,166 U.S.-resident singleton live births in the 2023 to 2024 National Vital Statistics System natality files. Low birthweight was defined as birthweight <2,500 g. Trajectories were never smoking, quitting before pregnancy, quitting during pregnancy, persistent smoking, or other/intermittent smoking. Modified Poisson regression estimated adjusted risk ratios (aRRs) and 95% confidence intervals controlling for birth year and maternal, pregnancy, and infant characteristics.

**Results:** Low-birthweight prevalence was 6.4% among never smokers, 7.7% among mothers who quit before pregnancy, 11.2% among those who quit during pregnancy, 14.6% among persistent smokers, and 10.2% among those with other/intermittent smoking. Compared with never smoking, adjusted risks were higher for quitting before pregnancy (aRR 1.06, 95% CI 1.03, 1.09), quitting during pregnancy (aRR 1.46, 95% CI 1.42, 1.51), persistent smoking (aRR 1.95, 95% CI 1.93, 1.98), and other/intermittent smoking (aRR 1.36, 95% CI 1.25, 1.49). Associations varied little by year (joint interaction P = 0.102).

**Conclusions:** Persistent smoking showed the strongest association with low birthweight. Because the outcome combines preterm delivery and impaired fetal growth, the estimates should not be interpreted as associations with fetal growth restriction alone.

**Synopsis:** 

**Study question:** How are maternal cigarette-smoking trajectories before and during pregnancy associated with low birthweight among U.S.-resident singleton live births in 2023–2024?

**What’s already known:** Maternal smoking during pregnancy is associated with low birthweight, and earlier smoking cessation is generally associated with more favourable birth outcomes.²–

**What this study adds:** Among 5,861,166 births, persistent smoking showed the strongest association with low birthweight (aRR 1.95, 95% CI 1.93, 1.98), followed by quitting during pregnancy (aRR 1.46, 95% CI 1.42, 1.51). Quitting before pregnancy showed only a small residual association (aRR 1.06, 95% CI 1.03, 1.09). Findings were consistent across 2023–2024 and support distinguishing smoking trajectories rather than using a simple binary exposure.

## Background

Maternal cigarette smoking remains a modifiable pregnancy exposure associated with impaired fetal growth and adverse birth outcomes. Although cigarette smoking during pregnancy has declined markedly in the United States, exposure has not disappeared. National Vital Statistics System (NVSS) data show that the proportion of women reporting cigarette smoking at any time during pregnancy fell from 3.0% in 2023 to 2.4% in 2024. Low birthweight, defined as birthweight below 2,500 g, nevertheless remained common nationally, accounting for 8.52% of all U.S. births in 2024.¹ These trends indicate that prenatal cigarette exposure warrants continued epidemiological attention despite declining prevalence.

Low birthweight is a pragmatic population indicator but is not aetiologically specific. It may result from shortened gestation, restricted fetal growth, or both, and these pathways have different risk-factor profiles. Associations with low birthweight therefore should not be interpreted as associations with fetal growth restriction alone.

The association between prenatal cigarette smoking and reduced birthweight is well established. A systematic review and meta-analysis of 55 cohort studies found a substantially higher occurrence of low birthweight among infants exposed to maternal smoking during pregnancy and reported evidence that associations were stronger with greater cigarette consumption and longer duration of smoking.² Importantly, duration of exposure may be as relevant as whether smoking occurred at all. A prospective multicentre cohort that classified women according to trimester of cessation found substantially less fetal growth restriction among women who stopped smoking during the first or second trimester than among women who continued smoking throughout pregnancy.³ These findings support analysing smoking as a time-varying behaviour rather than a single smoker/non-smoker indicator.

Evidence from U.S. populations also suggests that the timing of cessation matters. Xie and colleagues analysed birth records from 43 states and New York City and found more favourable birth outcomes among women who stopped smoking during pregnancy than among women who continued smoking. More recent evidence continues to support cessation during pregnancy. A 2025 meta-analysis of randomised trials of financial incentives for smoking cessation found higher mean birthweight among intervention groups, although estimates for low birthweight were less precise. A 2026 Cochrane review incorporating 127 studies and more than 47,000 participants found that psychosocial smoking-cessation interventions increased abstinence during late pregnancy and were associated with a lower risk of low birthweight (RR 0.82, 95% CI 0.73, 0.94).

Despite this evidence, several gaps remain. Many studies have evaluated smoking as a binary exposure, relied on intervention cohorts, or used older birth cohorts. Such approaches may obscure differences between women who stop smoking before conception, stop after pregnancy begins, continue smoking throughout pregnancy, or follow less consistent smoking patterns.

Contemporary national evidence distinguishing these trajectories is limited. The NVSS natality files are particularly suited to addressing this question because cigarette consumption is recorded separately for the three months before pregnancy and for each trimester. The NCHS also distinguishes women who quit before pregnancy from those who smoke during early pregnancy and stop before the third trimester.

A further methodological consideration concerns the choice of effect measure. For a common binary outcome such as low birthweight, odds ratios can diverge from relative risks and may be less intuitive for population-based interpretation. Accordingly, this study was designed to estimate risk ratios directly using modified Poisson regression with robust variance rather than logistic-regression odds ratios.

This study therefore examined the association between maternal cigarette-smoking trajectories and low birthweight among U.S.-resident singleton live births recorded in the 2023–2024 NVSS Natality Public-Use Files. Smoking was classified as never smoking, quitting before pregnancy, quitting during pregnancy, persistent smoking throughout pregnancy, or other/intermittent smoking. Secondary objectives examined any smoking during pregnancy, third-trimester smoking intensity, and consistency of trajectory associations across 2023 and 2024.

## Methods

### Study design and data source

We conducted a retrospective, population-based analysis of the 2023 and 2024 National Vital Statistics System (NVSS) Natality Public-Use Files maintained by the National Center for Health Statistics (NCHS). These files contain individual-level information from U.S. birth certificates and provide near-complete coverage of registered live births. The 2023 file contained 3,605,081

U.S. births, including 3,596,017 births to U.S. residents. The corresponding 2024 file contained 3,638,436 U.S. births, of which 3,628,934 were births to U.S. residents. Variable coding and edit specifications followed the accompanying 2023 and 2024 user guides., Because the files represent registered births rather than a probability sample, survey weights were not applied.

The study population was restricted to U.S.-resident singleton live births in 2023–2024. Births to foreign residents and multiple births were excluded. The primary complete-case analytic sample comprised **5,861,166 births**.

### Maternal cigarette-smoking trajectories

The primary exposure was maternal cigarette-smoking trajectory. NVSS records the average number of cigarettes smoked per day during the 3 months before pregnancy (CIG_0) and separately during the first (CIG_1), second (CIG_2), and third (CIG_3) trimesters. Values of 00– 97 represent the reported number of cigarettes smoked per day, 98 indicates 98 or more cigarettes per day, and 99 indicates unknown or not stated.

Five mutually exclusive trajectories were defined. Never smokers reported no cigarette use before pregnancy or during any trimester. Quit before pregnancy included women who smoked before pregnancy but reported no smoking during pregnancy. Quit during pregnancy included women who smoked during the first and/or second trimester but reported no smoking during the third trimester. Persistent smoking throughout pregnancy required reported smoking during all three trimesters. Remaining patterns were classified as other/intermittent smoking. Definitions followed NCHS guidance: smoking before but not during pregnancy was classified as quitting before pregnancy, and smoking only in the first and/or second trimester as quitting during pregnancy.

For pregnancies ending before 27 completed weeks, NCHS edits third-trimester cigarette use to unknown because the third trimester has not been fully observed. Consequently, the trajectory analysis required at least 27 completed weeks of gestation and known cigarette-use information for all four exposure periods. This restriction allowed smoking trajectories to be defined consistently but means that extremely preterm births were not represented in the primary trajectory analysis.

### Outcome

The primary outcome was low birthweight, defined as birthweight below 2,500 g. Birthweight was obtained from the detailed birthweight variable (DBWT), and records coded as not stated were treated as missing. NCHS and the International Classification of Diseases define low birthweight as birthweight below 2,500 g. The outcome variable was coded as 1 for low birthweight and 0 otherwise.

### Covariates

Covariates were specified before fitting the final models based on established relationships with maternal smoking and fetal growth and their consistent availability in the 2023 and 2024 natality files. The primary multivariable model adjusted for birth year; maternal age; race and Hispanic origin; maternal education; marital status; principal payment source for delivery; participation in the Special Supplemental Nutrition Program for Women, Infants, and Children (WIC); prepregnancy body mass index; live-birth order; prepregnancy diabetes; prepregnancy hypertension; previous preterm birth; and infant sex.

Reference categories were 2023, maternal age 25–29 years, non-Hispanic White, high school/GED education, married, private insurance, no WIC participation, normal prepregnancy BMI, first birth, no prepregnancy diabetes, no prepregnancy hypertension, no previous preterm birth, and female infant. Gestational diabetes, gestational hypertension, and prenatal-care variables were excluded from the primary adjustment set. Gestational age was not included because it may be downstream of maternal smoking and conditioning on it could block part of the association or introduce collider bias.

### Statistical analysis

Maternal and infant characteristics were summarised using counts and percentages. Low-birthweight prevalence was calculated overall, by birth year and across maternal smoking trajectories.

The primary association between maternal smoking trajectory and low birthweight was estimated using modified Poisson regression with a log link and heteroskedasticity-robust variance. This approach directly estimates risk ratios for binary outcomes and avoids interpreting logistic-regression odds ratios as relative risks. Crude risk ratios (RRs) and multivariable-adjusted risk ratios (aRRs) were reported with 95% confidence intervals. Never smoking served as the reference category.

The primary model was fitted using the feglm() function in the R package fixest, specifying family = poisson(link = "log") and heteroskedasticity-robust variance. The model estimated log risk ratios, which were exponentiated to obtain RRs and corresponding 95% confidence intervals.

### Missing data

The primary analysis used complete-case analysis. Records missing low birthweight, smoking trajectory, or any covariate included in the adjusted model were excluded. Unknown smoking responses were not recoded as non-smoking. The final primary modified-Poisson model included 5,861,166 births, of which 388,720 had low birthweight.

### Secondary analyses

A binary smoking analysis compared any cigarette smoking during pregnancy with no smoking during pregnancy using the same modified-Poisson specification and covariate set. The binary analysis included 5,861,166 births.

Third-trimester cigarette intensity was examined among 137,833 active third-trimester smokers. Cigarette consumption was classified as 1–5, 6–10, 11–20 or ≥21 cigarettes per day, with 1–5 cigarettes per day as the reference group. Modified Poisson regression with robust variance was again used to estimate adjusted RRs and 95% CIs.

Potential temporal variation was assessed by adding smoking-trajectory-by-birth-year interaction terms to the primary model. Ratios of RRs were estimated for 2024 relative to 2023, and the interaction terms were evaluated jointly using a four-degree-of-freedom Wald test.

### Ethics

The analysis used publicly available, de-identified NVSS public-use records. The investigators had no contact with mothers or infants and no access to names, addresses, medical-record numbers, or other direct personal identifiers. Accordingly, individual informed consent was not applicable to this secondary analysis.

## Results

### Study population and sample characteristics

The primary complete-case analysis included 5,861,166 U.S.-resident singleton live births in 2023–2024. Of these, 2,923,720 (49.9%) occurred in 2023 and 2,937,446 (50.1%) in 2024.

Overall, 388,720 infants had low birthweight, corresponding to a prevalence of 6.63%. The selection of births from the 2023 and 2024 NVSS natality files into the final analytic sample is summarised in Figure 1.

**Figure 1.**
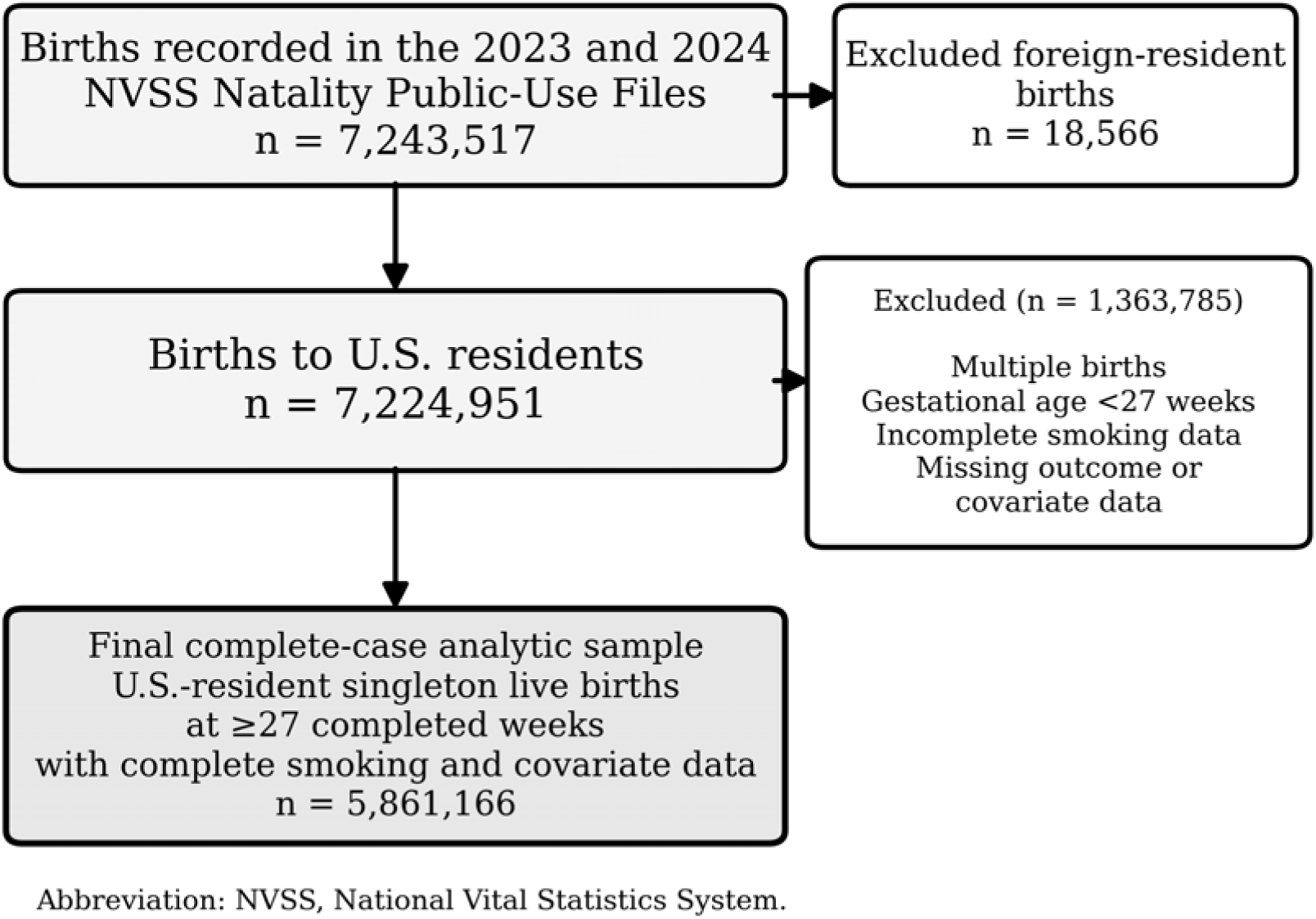
Selection of the primary analytic sample, United States, 2023-2024.

Characteristics of the analytic sample are presented in Table 1. Mothers aged 30–34 years constituted the largest age group (30.3%), followed by those aged 25–29 years (28.0%) and 20– 24 years (17.6%). Non-Hispanic White mothers accounted for 53.2% of births, Hispanic mothers for 24.0%, and non-Hispanic Black mothers for 14.2%. Approximately 60.3% of mothers were married. Private insurance was the principal payment source for 51.8% of births, while Medicaid accounted for 40.4%. WIC participation was reported for 30.9% of mothers. Prepregnancy diabetes, prepregnancy hypertension, and previous preterm birth were recorded for 1.28%, 3.34%, and 4.09% of births, respectively.

**Table 1.**
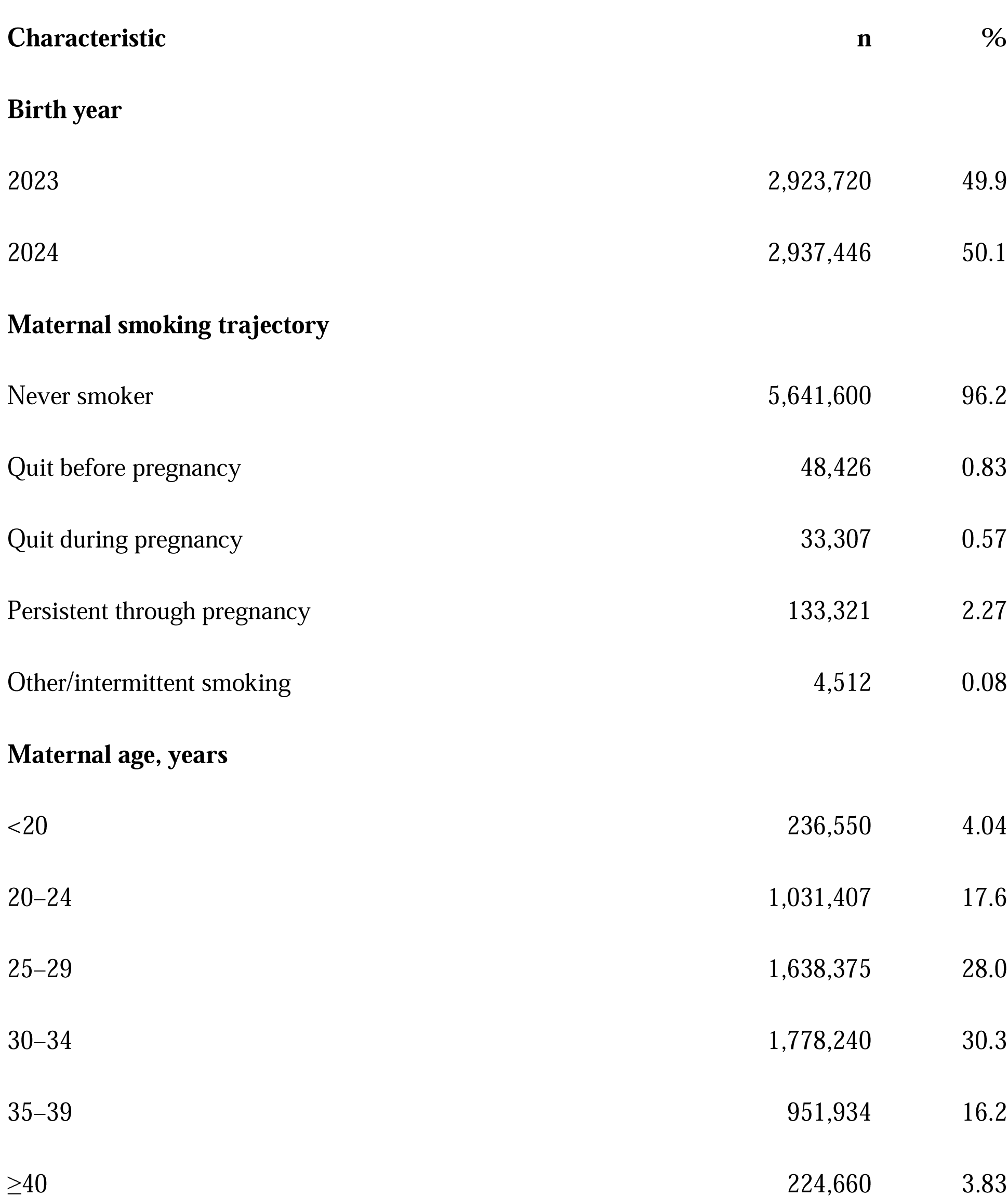
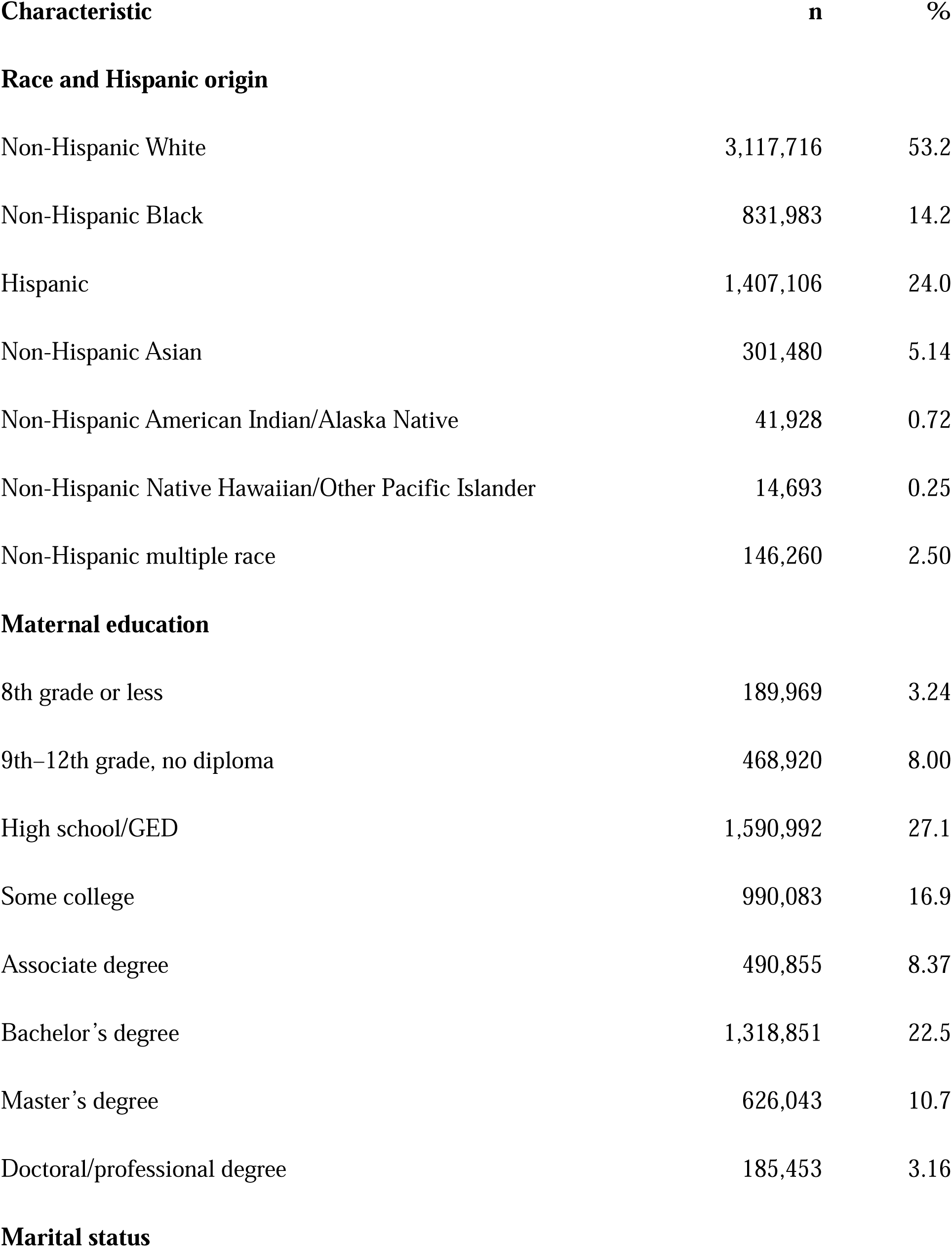
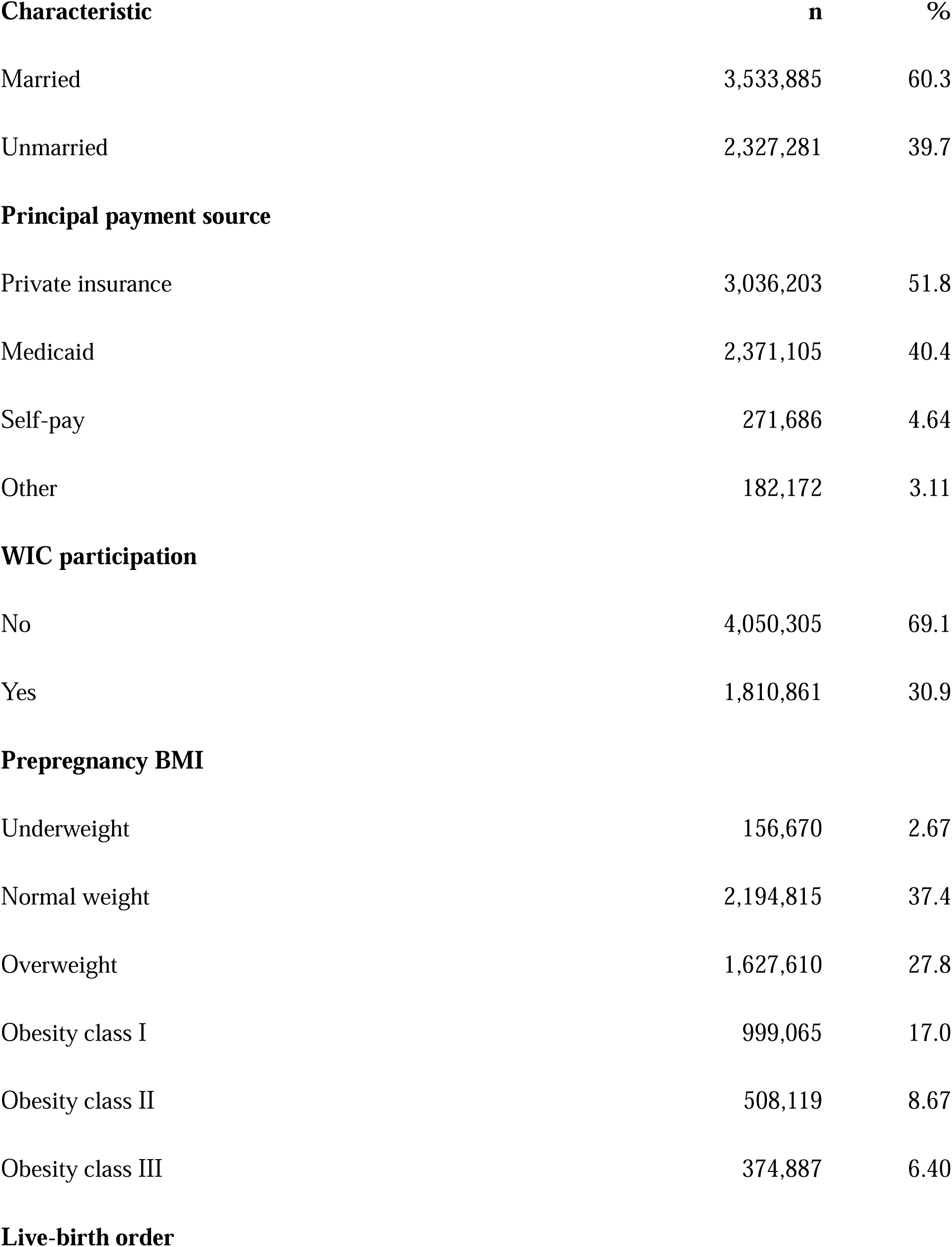
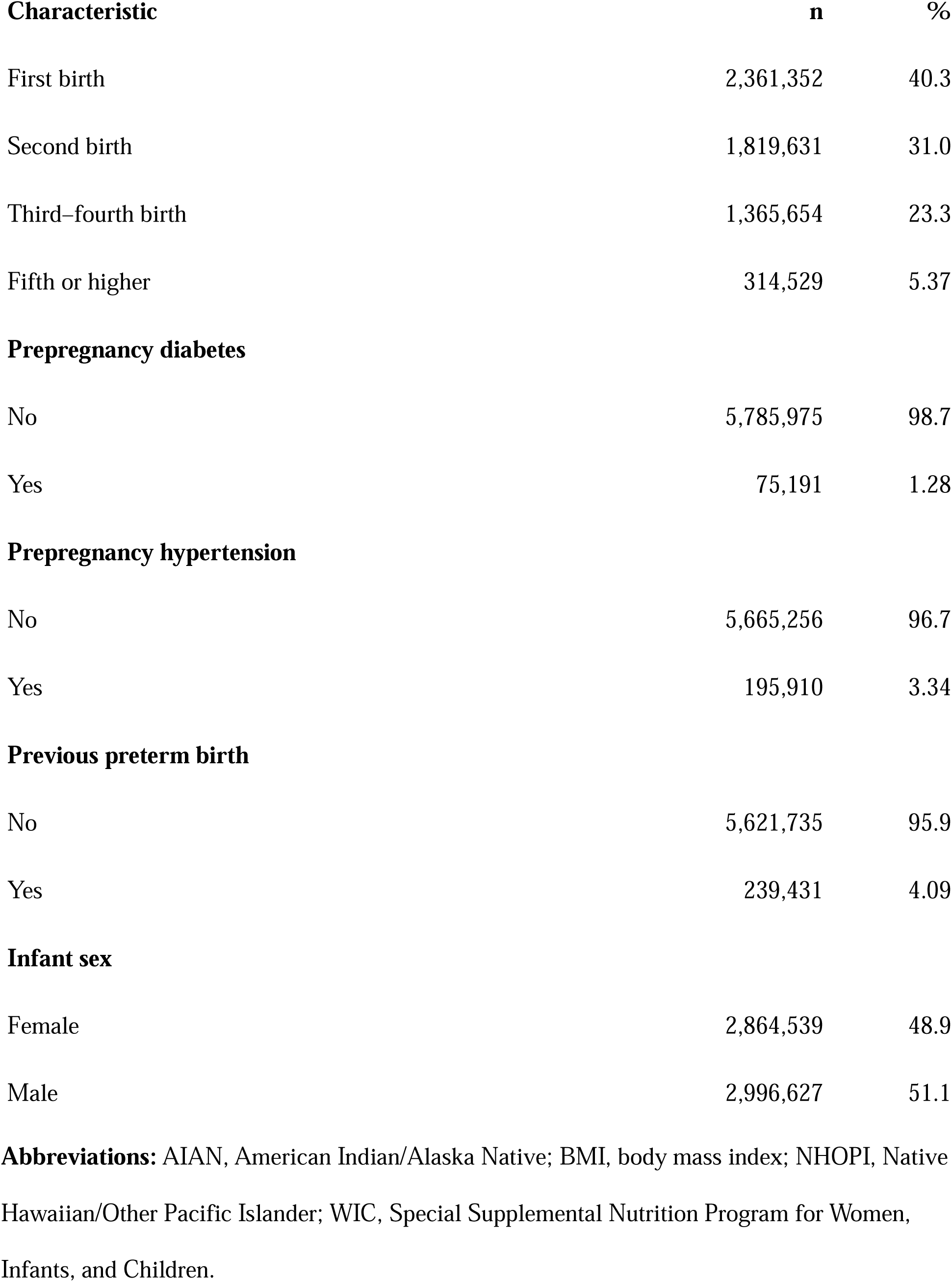
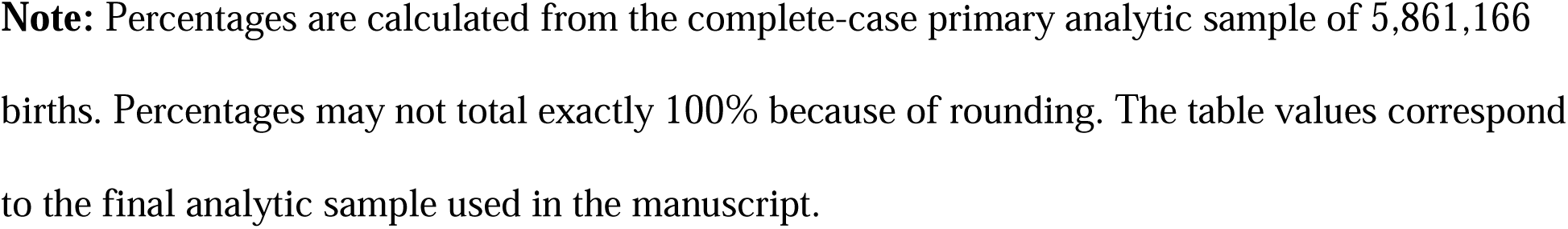
Characteristics of the primary analytic sample of U.S.-resident singleton live births, 2023-2024 (N = 5,861,166)

| **Characteristic** | **n** | **%** |
| --- | --- | --- |
| **Birth year** |  |  |
| 2023 | 2,923,720 | 49.9 |
| 2024 | 2,937,446 | 50.1 |
| **Maternal smoking trajectory** |  |  |
| Never smoker | 5,641,600 | 96.2 |
| Quit before pregnancy | 48,426 | 0.83 |
| Quit during pregnancy | 33,307 | 0.57 |
| Persistent through pregnancy | 133,321 | 2.27 |
| Other/intermittent smoking | 4,512 | 0.08 |
| **Maternal age, years** |  |  |
| <20 | 236,550 | 4.04 |
| 20–24 | 1,031,407 | 17.6 |
| 25–29 | 1,638,375 | 28.0 |
| 30–34 | 1,778,240 | 30.3 |
| 35–39 | 951,934 | 16.2 |
| ≥40 | 224,660 | 3.83 |
| **Race and Hispanic origin** |  |  |
| Non-Hispanic White | 3,117,716 | 53.2 |
| Non-Hispanic Black | 831,983 | 14.2 |
| Hispanic | 1,407,106 | 24.0 |
| Non-Hispanic Asian | 301,480 | 5.14 |
| Non-Hispanic American Indian/Alaska Native | 41,928 | 0.72 |
| Non-Hispanic Native Hawaiian/Other Pacific Islander | 14,693 | 0.25 |
| Non-Hispanic multiple race | 146,260 | 2.50 |
| **Maternal education** |  |  |
| 8th grade or less | 189,969 | 3.24 |
| 9th–12th grade, no diploma | 468,920 | 8.00 |
| High school/GED | 1,590,992 | 27.1 |
| Some college | 990,083 | 16.9 |
| Associate degree | 490,855 | 8.37 |
| Bachelor’s degree | 1,318,851 | 22.5 |
| Master’s degree | 626,043 | 10.7 |
| Doctoral/professional degree | 185,453 | 3.16 |
| **Marital status** |  |  |
| Married | 3,533,885 | 60.3 |
| Unmarried | 2,327,281 | 39.7 |
| **Principal payment source** |  |  |
| Private insurance | 3,036,203 | 51.8 |
| Medicaid | 2,371,105 | 40.4 |
| Self-pay | 271,686 | 4.64 |
| Other | 182,172 | 3.11 |
| **WIC participation** |  |  |
| No | 4,050,305 | 69.1 |
| Yes | 1,810,861 | 30.9 |
| **Prepregnancy BMI** |  |  |
| Underweight | 156,670 | 2.67 |
| Normal weight | 2,194,815 | 37.4 |
| Overweight | 1,627,610 | 27.8 |
| Obesity class I | 999,065 | 17.0 |
| Obesity class II | 508,119 | 8.67 |
| Obesity class III | 374,887 | 6.40 |
| **Live-birth order** |  |  |
| First birth | 2,361,352 | 40.3 |
| Second birth | 1,819,631 | 31.0 |
| Third–fourth birth | 1,365,654 | 23.3 |
| Fifth or higher | 314,529 | 5.37 |
| **Prepregnancy diabetes** |  |  |
| No | 5,785,975 | 98.7 |
| Yes | 75,191 | 1.28 |
| **Prepregnancy hypertension** |  |  |
| No | 5,665,256 | 96.7 |
| Yes | 195,910 | 3.34 |
| **Previous preterm birth** |  |  |
| No | 5,621,735 | 95.9 |
| Yes | 239,431 | 4.09 |
| **Infant sex** |  |  |
| Female | 2,864,539 | 48.9 |
| Male | 2,996,627 | 51.1 |

**Abbreviations:** AIAN, American Indian/Alaska Native; BMI, body mass index; NHOPI, Native Hawaiian/Other Pacific Islander; WIC, Special Supplemental Nutrition Program for Women, Infants, and Children.
**Note:** Percentages are calculated from the complete-case primary analytic sample of 5,861,166 births. Percentages may not total exactly 100% because of rounding. The table values correspond to the final analytic sample used in the manuscript.

### Maternal smoking trajectories and low birthweight

Most mothers were classified as never smokers, representing 5,641,600 births (96.2%). Persistent smoking throughout pregnancy was reported for 133,321 mothers (2.27%). A further 48,426 (0.83%) quit before pregnancy, 33,307 (0.57%) quit during pregnancy, and 4,512 (0.08%) had other or intermittent smoking patterns.

Low-birthweight prevalence varied across smoking trajectories (Table 2). Among never smokers, 361,272 births were low birthweight, corresponding to a prevalence of 6.4%. Prevalence was **7.7%** among mothers who quit before pregnancy, 11.2% among those who quit during pregnancy, 14.6% among persistent smokers, and 10.2% among mothers with other or intermittent smoking patterns. The corresponding unadjusted prevalence pattern is shown in Supplementary Figure 1.

**Table 2.** Maternal cigarette-smoking trajectory and risk of low birthweight among U.S.- resident singleton live births, 2023–2024.

| Smoking trajectory | N | Low birthweight, n (%) | Crude RR (95% CI) | Adjusted RR (95% CI) |
| --- | --- | --- | --- | --- |
| Never smoker | 5,641,600 | 361,272 (6.4) | 1.00 (Reference) | 1.00 (Reference) |
| Quit before pregnancy | 48,426 | 3,711 (7.7) | 1.20 (1.16, 1.23) | 1.06 (1.03, 1.09) |
| Quit during pregnancy | 33,307 | 3,747 (11.2) | 1.76 (1.70, 1.81) | 1.46 (1.42, 1.51) |
| Persistent through pregnancy | 133,321 | 19,532 (14.6) | 2.29 (2.26, 2.32) | 1.95 (1.93, 1.98) |
| Other/intermittent smoking | 4,512 | 458 (10.2) | 1.59 (1.45, 1.73) | 1.36 (1.25, 1.49) |

**Abbreviations:** CI, confidence interval; RR, risk ratio.
**Note:** Adjusted risk ratios were estimated using modified Poisson regression with heteroskedasticity-robust variance. The adjusted model included birth year, maternal age, race and Hispanic origin, education, marital status, payment source, WIC participation, prepregnancy body mass index, live-birth order, prepregnancy diabetes, prepregnancy hypertension, previous preterm birth, and infant sex. Never smoking was the reference category.

### Primary modified Poisson analysis

In crude analyses, the risk of low birthweight was higher for each smoking trajectory compared with never smoking. The crude RR was 1.20 (95% CI 1.16, 1.23) for quitting before pregnancy, 1.76 (95% CI 1.70, 1.81) for quitting during pregnancy, **2.29** (95% CI 2.26, 2.32) for persistent smoking, and 1.59 (95% CI 1.45, 1.73) for other or intermittent smoking.

After multivariable adjustment using modified Poisson regression with heteroskedasticity-robust variance, the estimates were attenuated but remained elevated. Compared with never smoking, mothers who quit before pregnancy had a modestly higher risk of low birthweight (aRR 1.06, 95% CI 1.03, 1.09). The association was stronger among mothers who quit during pregnancy (aRR 1.46, 95% CI 1.42, 1.51) and was greatest among mothers who smoked persistently throughout pregnancy (aRR 1.95, 95% CI 1.93, 1.98). Other or intermittent smoking was also associated with higher low-birthweight risk (aRR 1.36, 95% CI 1.25, 1.49) (Table 2).

Figure 2 shows the adjusted risk ratios and 95% confidence intervals for the four smoking trajectories. Persistent smoking showed the largest adjusted association with low birthweight.

**Figure 2.**
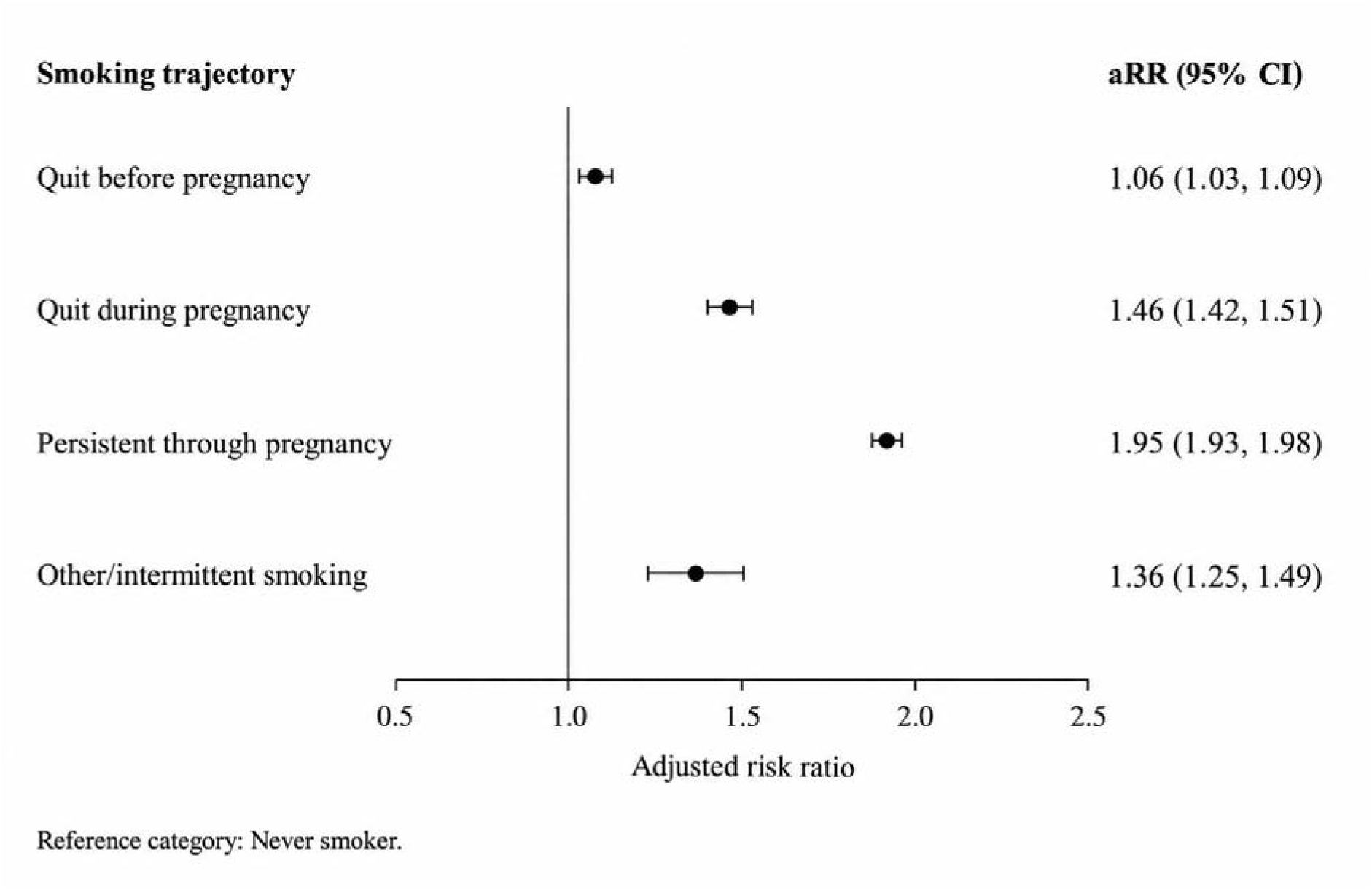
Adjusted risk ratios for low birthweight according to maternal cigarette-smoking trajectory, United States, 2023-2024.

### Secondary analyses

In the secondary binary-smoking analysis, any cigarette smoking during pregnancy was associated with a higher risk of low birthweight compared with no smoking after adjustment for the same covariates included in the primary model (aRR 1.83, 95% CI 1.81, 1.86).

Third-trimester cigarette intensity was evaluated among 137,833 active third-trimester smokers. Of these, 60,552 smoked 1–5 cigarettes per day, 49,723 smoked 6–10 cigarettes per day, 24,648 smoked 11–20 cigarettes per day, and 2,910 smoked ≥21 cigarettes per day. Compared with mothers smoking 1–5 cigarettes per day, the adjusted risk of low birthweight was higher among those smoking 6–10 cigarettes per day (aRR 1.08, 95% CI 1.05, 1.11), 11–20 cigarettes per day (aRR 1.14, 95% CI 1.10, 1.18), and ≥21 cigarettes per day (aRR 1.10, 95% CI 1.01, 1.20) (Table 3). The estimates did not show a strictly monotonic increase across smoking-intensity categories.

**Table 3.** Third-trimester cigarette intensity and adjusted risk of low birthweight among active smokers (N = 137,833)

| **Third-trimester cigarettes/day** | **N** | **Adjusted RR (95% CI)** |
| --- | --- | --- |
| 1–5 | 60,552 | 1.00 (Reference) |
| 6–10 | 49,723 | 1.08 (1.05, 1.11) |
| 11–20 | 24,648 | 1.14 (1.10, 1.18) |
| ≥21 | 2,910 | 1.10 (1.01, 1.20) |

**Abbreviations:** CI, confidence interval; RR, risk ratio.
**Note:** Adjusted risk ratios were estimated using modified Poisson regression with heteroskedasticity-robust variance among mothers who reported active cigarette smoking during the third trimester. Models adjusted for birth year, maternal age, race and Hispanic origin, education, marital status, payment source, WIC participation, prepregnancy body mass index, live-birth order, prepregnancy diabetes, prepregnancy hypertension, previous preterm birth, and infant sex. Smoking 1–5 cigarettes/day was the reference category. The estimates did not show a strictly monotonic increase across higher smoking-intensity categories.

### Variation by birth year

Smoking-trajectory associations were broadly similar in 2023 and 2024. Ratios of RRs comparing 2024 with 2023 were 1.07 (95% CI 1.01, 1.14) for quitting before pregnancy, 1.02 (95% CI 0.96, 1.08) for quitting during pregnancy, 1.00 (95% CI 0.98, 1.03) for persistent smoking, and 1.15 (95% CI 0.97, 1.36) for other or intermittent smoking. The joint four-degree-of-freedom Wald test provided little evidence of overall heterogeneity in the trajectory associations by birth year (P = 0.102).

### Comment

#### Principal findings

In this population-based analysis of 5,861,166 U.S.-resident singleton live births in 2023–2024, associations with low birthweight varied across maternal cigarette-smoking trajectories. Low-birthweight prevalence was 6.4% among never smokers, 7.7% among mothers who quit before pregnancy, 11.2% among those who quit during pregnancy, 14.6% among persistent smokers, and 10.2% among those with other or intermittent smoking patterns (Table 2; Supplementary Figure 1). After multivariable adjustment using modified Poisson regression with heteroskedasticity-robust variance, persistent smoking had the largest association with low birthweight (aRR 1.95, 95% CI 1.93, 1.98), followed by quitting during pregnancy (aRR 1.46, 95% CI 1.42, 1.51), other or intermittent smoking (aRR 1.36, 95% CI 1.25, 1.49), and quitting before pregnancy (aRR 1.06, 95% CI 1.03, 1.09) (Table 2; Figure 2).

The secondary analyses reinforced the primary findings. Any cigarette smoking during pregnancy was associated with higher low-birthweight risk than no smoking (aRR 1.83, 95% CI 1.81, 1.86). Among 137,833 active third-trimester smokers, adjusted risk was higher for 6–10 cigarettes/day (aRR 1.08, 95% CI 1.05, 1.11), 11–20 cigarettes/day (aRR 1.14, 95% CI 1.10, 1.18), and ≥21 cigarettes/day (aRR 1.10, 95% CI 1.01, 1.20) compared with 1–5 cigarettes/day (Table 3). The intensity estimates did not form a strictly monotonic pattern. Associations were broadly consistent across 2023 and 2024, with little evidence of overall trajectory-by-year heterogeneity (joint interaction *P* = 0.102).

#### Strengths of the study

A major strength is the size, recency, and national coverage of the NVSS natality data. The analytic cohort provided sufficient precision to distinguish smoking patterns that would be difficult to examine reliably in smaller studies. Rather than reducing cigarette exposure to a smoker/non-smoker indicator, the study used reported consumption before pregnancy and during each trimester to distinguish quitting before pregnancy, quitting during pregnancy, persistent smoking, and other or intermittent patterns. Figure 1 summarises the cohort-selection process, and Table 1 provides the characteristics of the final analytic population.

The analytic approach is another strength. Modified Poisson regression with a log link and robust variance produced risk ratios directly, avoiding interpretation of logistic-regression odds ratios as risks. Secondary binary-smoking, intensity, and temporal-interaction analyses examined the consistency of the primary findings from several perspectives.

#### Limitations of the data

The primary limitation is that low birthweight is a heterogeneous endpoint that may reflect preterm delivery, restricted fetal growth, or both. These pathways have distinct epidemiological profiles, and this study did not estimate associations separately for preterm birth and small for gestational age. The results therefore apply to low birthweight as defined and should not be interpreted as estimates of fetal growth restriction.

Several limitations should temper interpretation. Cigarette use recorded on the birth certificate is self-reported and may be underreported or misclassified. In a validation study comparing U.S. birth-certificate smoking information with cotinine measurements, birth certificates identified most but not all prenatal smokers, and the correlation between reported third-trimester cigarettes/day and cotinine was moderate.^10^ Such measurement error could reduce contrasts between smoking groups and may partly contribute to the non-monotonic pattern observed in the highest smoking-intensity categories.

Trajectory classification also required gestational age of at least 27 completed weeks because third-trimester cigarette information is not fully observable for earlier deliveries. Extremely preterm births, which carry a high probability of low birthweight, were therefore not represented in the trajectory analysis. The findings should not be generalised to births before 27 weeks, and this restriction partly explains why low-birthweight prevalence in the analytic population was lower than prevalence in an unrestricted national birth population.

Complete-case analysis may also have introduced selection if missing exposure or covariate information was related to both smoking and birthweight. The public-use files do not permit repeated births to the same mother to be identified, and residual confounding remains possible from factors that were unavailable or incompletely measured, including second-hand smoke, other nicotine products, diet, psychosocial stress, detailed socioeconomic circumstances, and other substance use. The other/intermittent-smoking group was small and heterogeneous, while the ≥21-cigarettes/day category contained only 2,910 mothers, resulting in less precise estimation.

#### Interpretation

The findings are consistent with evidence that both the duration and timing of smoking exposure are associated with birthweight-related outcomes. Di and colleagues’ meta-analysis of 55 cohort studies found a strong association between smoking during pregnancy and low birthweight, with stronger associations for longer smoking duration and greater cigarette consumption.^2^ Xie and colleagues, analysing U.S. PRAMS data from 2012–2017, likewise reported less favourable birth outcomes among women who continued smoking than among those who stopped.^4^ Prospective biomarker-supported research has also reported more favourable fetal-growth outcomes when smoking cessation occurs earlier in pregnancy.^3^

The present study extends this literature by showing the same broad pattern in contemporary 2023–2024 U.S. birth records while directly estimating adjusted risk ratios. Persistent exposure had the strongest association, whereas the estimate among mothers who quit before pregnancy was close to the null. The small residual association in the quit-before-pregnancy group should not be interpreted as evidence that prior smoking independently causes low birthweight; residual socioeconomic or behavioural differences, cumulative smoking history, and exposure misclassification remain plausible explanations.

Several biological pathways could contribute to the observed pattern. Cigarette-smoke constituents may impair placental development and function by altering uteroplacental circulation, increasing oxidative stress, causing cellular injury, and restricting oxygen and nutrient delivery. Hoch and colleagues found increased DNA damage and evidence of disturbed early placental development in first-trimester placental tissue from smokers.^11^ These placental effects provide biologically plausible pathways through which continued smoking could contribute to lower birthweight.

Maternal smoking may contribute to low birthweight through pathways involving gestational duration, fetal growth, or both. Because gestational age may be downstream of smoking and conditioning on it can introduce collider bias, no gestational-age-adjusted estimate is presented. The reported associations concern low birthweight as recorded and should not be interpreted as associations with fetal growth restriction or preterm birth separately.

The findings also have practical implications for smoking cessation. The substantially lower estimate among mothers who stopped smoking during pregnancy than among persistent smokers is consistent with benefit from stopping after conception, although this observational analysis cannot quantify a causal cessation effect. Intervention evidence strengthens this interpretation. A 2025 meta-analysis of randomised trials found higher mean birthweight when financial incentives were offered for smoking cessation,^5^ while the 2026 Cochrane review of 127 studies concluded that psychosocial interventions improve late-pregnancy abstinence and may reduce low birthweight.^6^ These findings support repeated opportunities for smoking-cessation assistance before conception and throughout prenatal care.

## Conclusions

Persistent cigarette smoking throughout pregnancy was associated with the highest risk of low birthweight. Mothers who quit during pregnancy had a lower estimated risk than persistent smokers but remained at higher risk than never smokers, whereas quitting before pregnancy showed only a small residual association. The consistency of estimates across 2023 and 2024 supports the contemporary relevance of these findings. Low birthweight is a heterogeneous endpoint, and the results should not be interpreted as associations with fetal growth restriction or preterm birth separately. Because this study is observational and relies on birth-certificate smoking reports, the estimates describe associations rather than causal effects.

## Data Availability Statement

The data that support the findings of this study are publicly available from the National Center for Health Statistics (NCHS) through the Vital Statistics Online Data Portal. This study used the 2023 and 2024 U.S. Natality Public-Use Files: https://www.cdc.gov/nchs/data_access/vitalstatsonline.htm.

## Data Availability

All data analyzed in this study are publicly available from the Centers for Disease Control and Prevention, National Center for Health Statistics, through the 2023 and 2024 National Vital Statistics System Natality Public Use Data Files: https://www.cdc.gov/NCHS/data_access/VitalStatsOnline.htm. The analytic code is available from the corresponding author upon reasonable request.

---

**Supplementary Figure 1.**
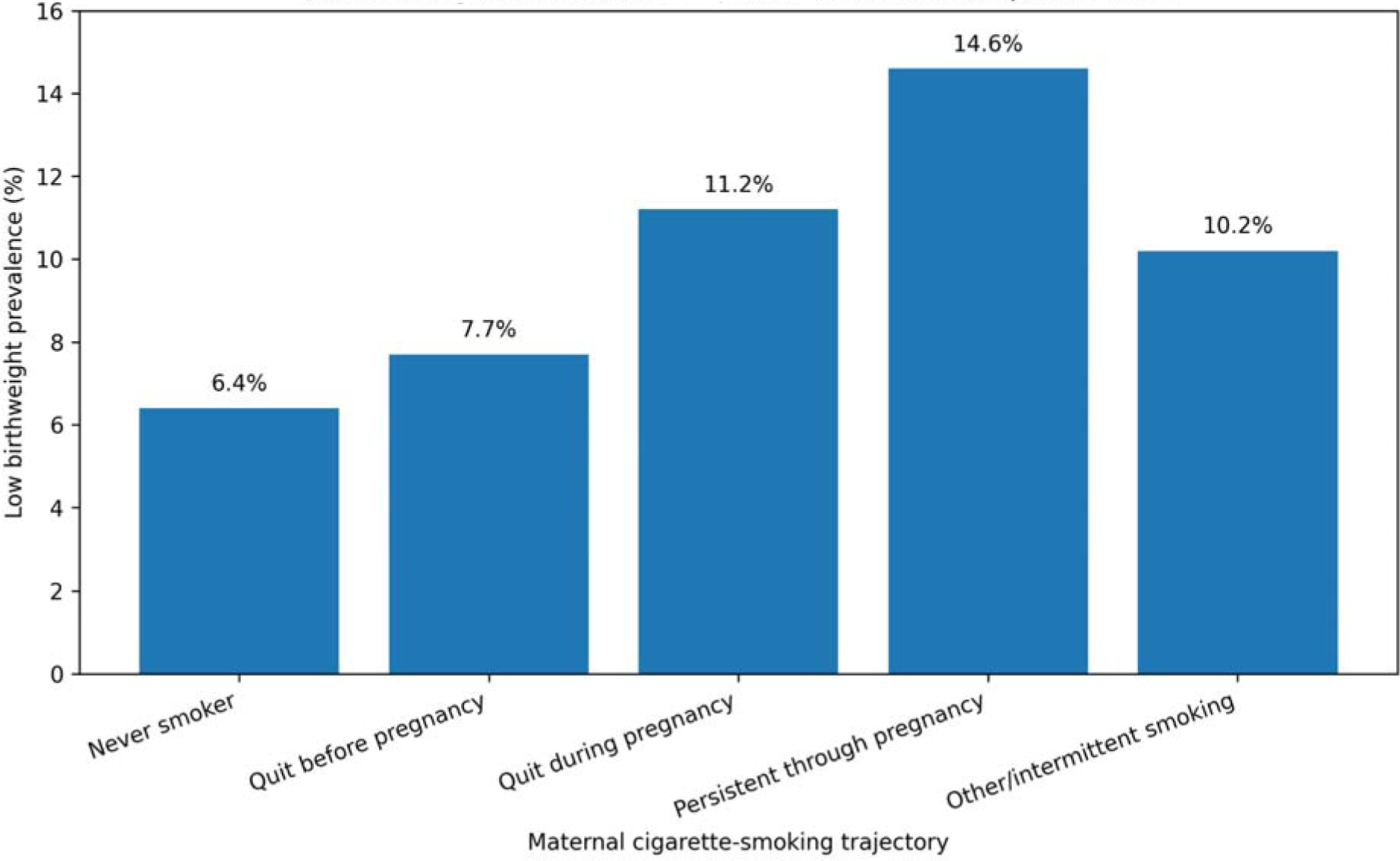
Prevalence of low birthweight according to maternal cigarette-smoking trajectory, United States, 2023-2024.

