## Supplementary figures and images for "Maternal Cigarette Smoking Trajectories and Risk of Low Birthweight in the United States: A Population-Based Analysis of 2023 to 2024 Natality Data"

### Supplemental Data 1

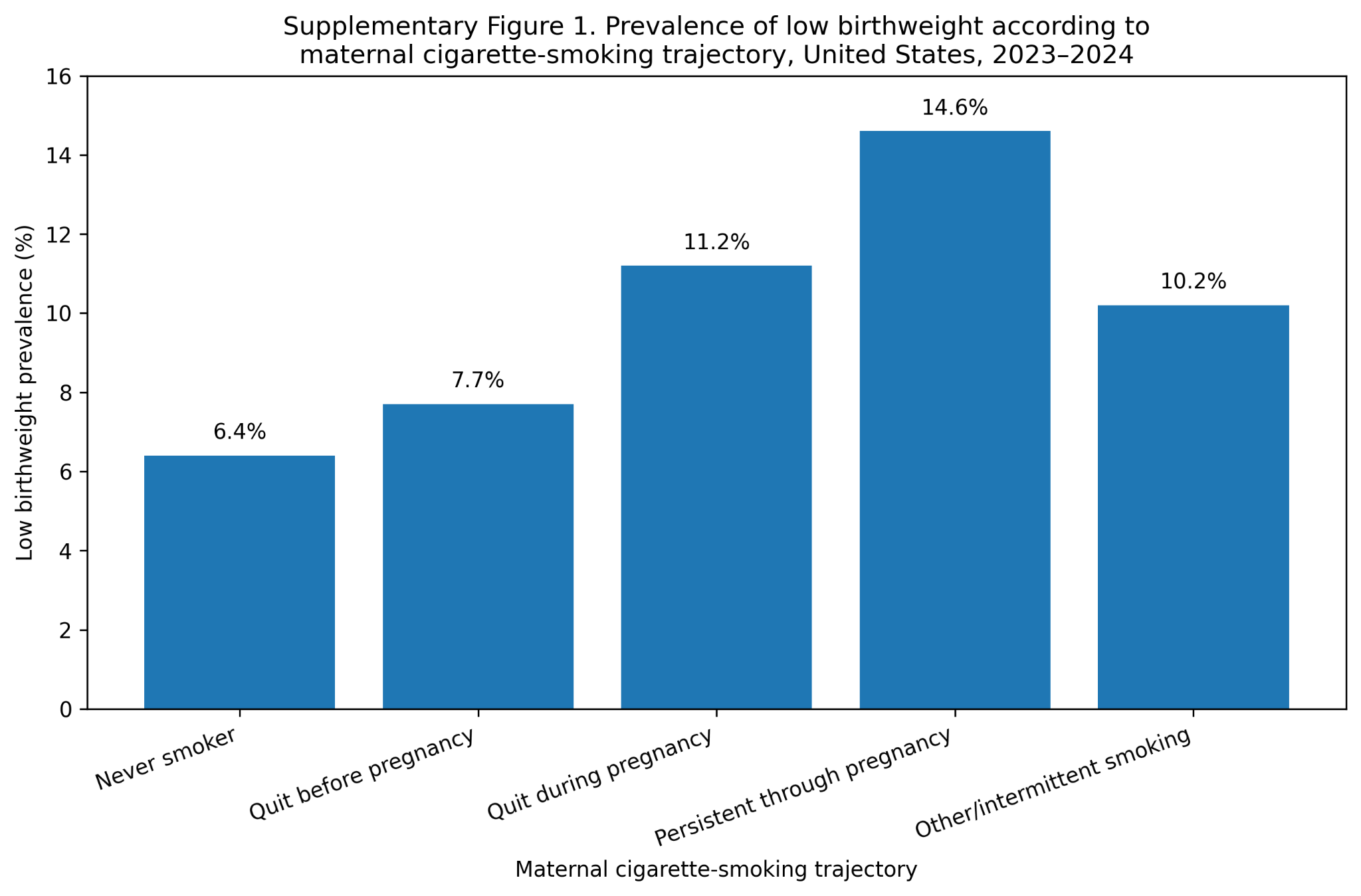
